# Association between gastric rhythm and gastroesophageal reflux defined by simultaneous body surface gastric mapping and 24-hour pH testing

**DOI:** 10.1101/2025.07.28.25332302

**Authors:** William Xu, Sam Simmonds, Daphne Foong, Sameer Bhat, Chris Varghese, Christopher N Andrews, Gabriel Schamberg, Armen Gharibans, Thomas L. Abell, David Rowbotham, Vincent Ho, Stefan Calder, Gregory O’Grady

## Abstract

**Background:** Abnormal gastric myoelectrical function may contribute to gastroesophageal reflux disease (GERD). We assessed if myoelectrical abnormalities measured using body surface gastric mapping were correlated with reflux measured by 24 hr pH testing, and symptom severity.

**Methods:** Gastric Alimetry^®^ was performed simultaneously on patients undergoing 24 hr pH testing for investigation of reflux symptoms, with a standardised 4.5 hr test and validated symptom logging. Data were segmented into 15-minute epochs. Correlations between myoelectric activity, reflux events, and symptoms were assessed, including temporal correlations adjusted for repeated measures.

**Results:** Forty subjects were recruited (mean age 46.5 years, 60% female): 20 undergoing pH testing (12 with GERD and 8 symptomatic patients without), and 20 controls. GERD patients displayed less stable Gastric Alimetry® Rhythm-Index (GA-RI) compared with controls (p=0.011), but not with non-GERD patients (p=0.605). Decreasing GA-RI was associated with esophageal acid exposure (DeMeester score; r=-0.46, p=0.042). Periods of decreased GA-RI were not temporally correlated with reflux (r=0.08, p=0.182), or heartburn severity (r=0.04, p=0.309), but were correlated with nausea (r=-0.22, p<0.001) and excessive fullness (r=-0.28, p<0.001).

**Conclusion:** Gastric rhythm instability is associated with increased symptom severity and overall acid exposure in GERD patients, although no temporal link to heartburn was found. Reduced rhythm stability was temporally associated with increased nausea and fullness. GA-RI offers an emerging biomarker of gastric dysfunction in patients with GERD symptomatology.

**Study highlights:** *What is known:*

● Heartburn is common and often medically refractory
● Gastric conduction and motility abnormalities may contribute to symptoms but the temporal relationship is unknown

*What is new here:*

● Gastric rhythm abnormalities measured by Gastric Alimetry® are correlated with increasing reflux burden and symptom severity
● There is no temporal association between gastric rhythm and reflux events
● Gastric rhythm abnormalities may predispose patients to worse reflux, but there is no direct temporal correlation

## Introduction

Gastroesophageal reflux disease (GERD) affects 10% – 20% of the adult population in the Western world,^1^ and is associated with substantial decreases in health-related quality of life especially in cases of medically-refractory symptoms.^2^ The predominant pathogenic mechanism is acid in the proximal stomach,^2^ and reflux into the esophagus during dysfunctional lower esophageal sphincter relaxation.^3,4^ Esophageal abnormalities such as hiatal hernias, impaired peristalsis, and hypersensitivity also contribute.^2^

While GERD is viewed as primarily an esophageal disorder, impaired proximal stomach emptying,^5^ and impaired fundic accommodation may also be significant.^6^ Furthermore, gastric dysrhythmias have been more commonly observed in patients with GERD than those without,^7,8^ particularly in those with regurgitation symptoms.^8^ Extant literature have relied on legacy electrogastrography (EGG) methods which are limited by noise contamination and a lack of reproducible biomarkers.^9^ It remains unclear whether gastric dysrhythmias in GERD are a primary causative mechanism, exacerbating factor, or feature of concurrent gastric dysmotility.

Body Surface Gastric Mapping (BSGM) using Gastric Alimetry^®^ (Auckland, New Zealand) is a novel, non-invasive diagnostic technique without the limitations of legacy EGG methods.^10–12^ Gastric Alimetry® employs a dense array of electrodes at the epigastrium to measure gastric myoelectrical activity simultaneously with upper gastrointestinal symptom logging, and has been extensively validated.^10,13,14^

The aim of this study was to perform simultaneous 24 hr pH monitoring and Gastric Alimetry^®^. We hypothesised that gastric dysrhythmia would correlate with symptom severity, and temporally correlate with reflux measured on 24 hr pH-monitoring and with symptoms.

## Methods

Ethical approval for reporting was obtained from the Auckland Health Ethics Research Ethics Committee and Western Sydney Human Research Ethics Committee (ref: AH1130, H15157). All patients provided informed consent.

Consecutive adult patients referred for 24 hr pH-monitoring testing in Auckland, New Zealand and Western Sydney, Australia were approached for recruitment. Referred patients were evaluated for upper gastrointestinal symptoms consistent with suspected GERD either for pre-surgical workup, refractory symptoms, or for diagnostic clarification. Patients were excluded if they had previous upper gastrointestinal surgery, active gastrointestinal malignancy, active neurogenic or endocrine disorders affecting gastric motility (*e.g.*, multiple sclerosis, scleroderma, Parkinson’s disease, hyperthyroidism), a current pregnancy, or cognitive impairment. Routine BSGM Gastric Alimetry® test exclusions were also applied (BMI > 35, fragile or damaged epigastric skin, unable to remain still, or major allergy to adhesives).^15^ If patients were on proton pump inhibitor (PPI) medication, PPI medications were withheld at the discretion of the investigating gastroenterologist.

### Study procedure

All patients completed 24 hr pH monitoring tests, with catheter placement at the discretion of the gastroenterologist. Patients in Auckland were assessed using a pH impedance monitor (ZandorpH™, Laborie, Australia), while those in Sydney were assessed using a pH monitor (AccuView™ pH/pH-Z, Medtronic, USA).

During the 24 hr pH monitoring window, all patients underwent a 4.5 hr period of simultaneous BSGM (Gastric Alimetry® , New Zealand), as previously described (**Supplementary Fig. 1**).^10^ The Gastric Alimetry® Device consists of a high-resolution flexible electrode array (8 × 8 electrodes; 20 mm spacing; 196 cm^2^), wearable reader, validated iOS app for symptom logging, and cloud-based reporting platform.^12^ Patients withheld medications affecting GI motility including prokinetics and fasted for at least 6 hr before undergoing a 30 min baseline recording, followed by a standardized meal consisting of Ensure (232 kcal, 250 mL; Abbott Nutrition, USA) and an energy bar or similar (250 kcal, 5 g fat, 45 g carbohydrate, 10 g protein, 7 g fibre; Clif Bar & Company, USA), and a 4 hr postprandial recording. Patients were encouraged to eat as much as possible to stimulate symptoms.^10^ The timing of BSGM recordings was pragmatically scheduled relative to the pH-test; BSGM was conducted in the morning for patients whose pH-monitoring began in the morning, and on the following morning for those whose pH-testing began in the afternoon.

### Symptom assessment

Baseline symptom-severity and quality-of-life reporting were completed with the Patient Assessment of Upper Gastrointestinal Disorders-Symptom Severity Index (PAGI-SYM), Gastroparesis Cardinal Symptom Index (GCSI), and Patient Assessment of Upper Gastrointestinal Disorders-Quality of Life (PAGI-QOL) instruments.^16,17^

During BSGM testing, patients simultaneously logged their gastroduodenal symptoms profiles in the validated Gastric Alimetry® App, comprising visual scales for nausea, bloating, upper gut pain, heartburn, stomach burn and excessive fullness (0-10).^14^ Total Symptom Burden was calculated as the sum of each participant’s mean postprandial symptom scores including early satiation.^14^

### Patient cohorts and definitions

Participants undergoing pH monitoring were age-, sex-, and BMI-matched to controls.^18^ Control subjects were excluded if they had active gastrointestinal symptoms, met Rome IV criteria, were taking medications altering gastrointestinal motility, consumed regular cannabis, or had > 50% of the test duration marked as artifacts, as per the Gastric Alimetry® Instructions for Use.

Patients were stratified into those with and without a diagnosis of GERD based on physiological testing. A positive diagnosis of GERD was defined as either a positive pH test (either an indicative DeMeester score or significant symptom association interpreted by a specialist gastroenterologist), or evidence of reflux esophagitis on previous endoscopy.^2,8^

### Data Processing

The following metrics of gastric myoelectrical function were analysed:

● Principal Gastric Frequency (PGF): frequency of stable gastric slow wave activity.

High PGF (> 3.35 cpm) has been associated with possible vagal neuropathy or injury.^20,21^

● Gastric Alimetry Rhythm Index (GA-RI): measure of gastric rhythm stability defined as the percentage of power within the gastric frequency range. Low GA-RI (< 0.25) suggestive of possible neuromuscular dysfunction.
● BMI-Adjusted Amplitude: amplitude of the measured signal adjusted for BMI via a multiplicative regression model.^13^
● fed:fasted amplitude ratio (ff-AR): maximal 1 hr postprandial amplitude divided by the preprandial amplitude.^13^

Meal response ratios (MRR) were calculated for each subject as the ratio of BMI-adjusted amplitude in the first 2 hr divided by the last 2 hr of BSGM recording, to quantify timing of maximal post-prandial amplitude.^22^ BSGM metrics were compared to quantified parameters from the pH recordings: number of acidic reflux episodes (pH < 4), length of each reflux episode(s), percentage of time spent in reflux (over the entire 24 hr, and during the period of simultaneous BSGM; %).

Overall DeMeester score was calculated as a function of acid exposure across 24 hr of ambulatory recording.^23^ pH recordings were also synchronised and truncated to match BSGM recordings, and post-prandial BSGM and reflux data were then segregated into 15 min epochs to increase the temporal resolution of comparisons across this period.

### Statistical analysis

Data are reported as the mean ± standard deviation (SD) or median and interquartile range (IQR). The chi-squared test of independence was used to compare categorical variables.

Continuous independent variables were compared using mixed effect models and the Mann-Whitney U test. Further pairwise comparisons were corrected for by the Tukey post-hoc test when data were compared across more than two groups. Correlations between demographic data, BSGM metrics, patient symptom symptoms and quality of life were assessed using Pearson correlation co-efficient (*r*). Mean-centred BSGM metrics and symptom scores were calculated to compare the impact of intra-individual changes on associations between metrics. Sensitivity analysis was conducted to determine the effect of withholding PPI drugs during testing.

A statistical significance threshold *p* < 0.05 was used for all tests. Analyses were performed in R version 4.2.0 (R Foundation for Statistical Computing, Austria) and Python version 3.9 (Python Software Foundation, USA).

## Results

A total of 20 patients undergoing pH testing for upper gastrointestinal symptoms (mean age 46.5 years, mean BMI 26.8 kg/m², 60 % female; **Table 1**) were recruited, alongside 20 matched controls (mean age 46.1 years, mean BMI 25.5 kg/m², 55% female; **Table 1**).

**Table 1.** Overall patient and control characteristics, presented as mean and standard deviation.

| Variable | Control | non-GERD | GERD | Total |
| --- | --- | --- | --- | --- |
| Number (% of total) | 20 (50.0) | 8 (20.0) | 12 (30.0) | 40 (100.0) |
| Age (years) | 46.1 ± 15.9 | 40.1 ± 12.0 | 48.7 ± 19.4 | 45.7 ± 16.3 |
| Number of Females (% of group) | 11 (55.0) | 6 (75.0) | 7 (58.3) | 24 (60.0) |
| BMI (kg•m<sup>-2</sup>) | 25.5 ± 3.5 | 27.1 ± 6.6 | 26.5 ± 3.5 | 26.1 ± 4.2 |
| Anxiety | 0 (0.0) | 3 (37.5) | 3 (25.0) | 6 (15.0) |
| Depression | 0 (0.0) | 3 (37.5) | 2 (16.7) | 5 (12.5) |
| Irritable bowel syndrome | 1 (5.0) | 3 (37.5) | 1 (8.3) | 5 (12.5) |

Overall, 12 patients had a confirmed diagnosis of GERD (7 patients had positive 24 hr pH tests, 2 patients had previous reflux esophagitis on gastroduodenoscopy, and 3 patients had both), while 8 symptomatic patients had no evidence of GERD (hereon referred to as non-GERD patients). Demographics and co-morbidities are described in **Table 1**.

### Clinical Investigations and Symptom Burden

GERD patients had a greater number of reflux events compared with non-GERD patients over the 24 hr (74.5 vs 30.5 events, p = 0.015; **Table 2**). Similarly, GERD patients had greater DeMeester scores than non-GERD patients (20.5 versus 3.8, p = 0.003), and spent greater overall time in reflux (5.4% vs 0.8%, p = 0.004; **Table 2**). GERD and non-GERD patients had similar frequency of hiatal hernias (*p* = 0.313,**Table 2**).

**Table 2:** Clinical investigation results between patients with and without gastro-esophageal reflux disease.

| Variable |  | non-GERD | GERD | Total | p |
| --- | --- | --- | --- | --- | --- |
| **Total N (%)** |  | **8 (40.0)** | **12 (60.0)** | **20** |  |
| PPI Medication withheld for pH test | Not withheld | 2 (25.0) | 4 (33.3) | 6 (30.0) | 1.000 |
|  | Withheld/not on PPI | 6 (75.0) | 8 (66.7) | 14 (70.0) |  |
| DeMeester score | Median (IQR) | 3.8 (3.1 to 5.2) | 20.5 (9.4 to 28.0) | 6.9 (3.9 to 21.5) | **0.003** |
| Prev. hiatal hernia on gastroscopy (%) |  | 2 (25.0) | 7 (58.3) | 9 (45.0) | 0.313 |
| % time in acid exposure over pH test | Median (IQR) | 0.8 (0.7 to 1.0) | 5.3 (2.6 to 7.5) | 1.4 (0.9 to 5.6) | **0.004** |
| No. of reflux episodes over pH test | Median (IQR) | 30.5 (20.5 to 45.5) | 74.5 (50.5 to 128.2) | 55.0 (34.8 to 82.5) | **0.015** |
| Prev manometry - LES resting pressure | Low LES resting pressure | 1 (12.5) | 3 (25.0) | 4 (20.0) | 0.757 |
|  | Normal LES resting pressure | 5 (62.5) | 7 (58.3) | 12 (60.0) |  |
|  | No previous manometry | 2 (25.0) | 2 (16.7) | 4 (20.0) |  |

Both GERD and non-GERD patients had higher overall gastrointestinal symptom burden than controls (all *p* > 0.05, **Supplementary Table 1**). GERD patients had significantly higher heartburn symptom burden measured by PAGI-SYM subscale scores compared to non-GERD patients (2.7 vs 0.8, *p* = 0.025, **Supplementary Table 1**) but comparable symptom severity in other symptom subscales (all *p* > 0.5, **Supplementary Table 1**).

### Body Surface Gastric Mapping

Eighteen patients (90%) had normal spectral phenotypes defined by Gastric Alimetry® , with quantified values within the reference range (*e.g.*, GA-RI ≥ 0.25), though two GERD patients (10%) were classified as dysrhythmic (GA-RI < 0.25), both with hiatal hernias. Example spectrograms are displayed in **Fig. 1** and summary spectrograms are displayed in **Supplementary Fig. 2** GERD patients displayed lower overall GA-RI, indicating lower gastric rhythm stability, compared to controls (0.43 ± 0.16 vs. 0.61 ± 0.20; *p* = 0.029; **Table 3**). GA-RI was not statistically different between GERD and non-GERD patients when comparing overall values (0.43 ± 0.16 vs. 0.52 ± 0.08; *p* = 0.208; **Table 3**). However, post-prandial data assessed in 15-minute epochs demonstrated that GERD patients had a lower distribution of GA-RI values compared with non-GERD patients (0.46 ± 0.23 vs. 0.60 ± 0.21; *p* < 0.001; **Fig. 2**).

**Fig. 1.**
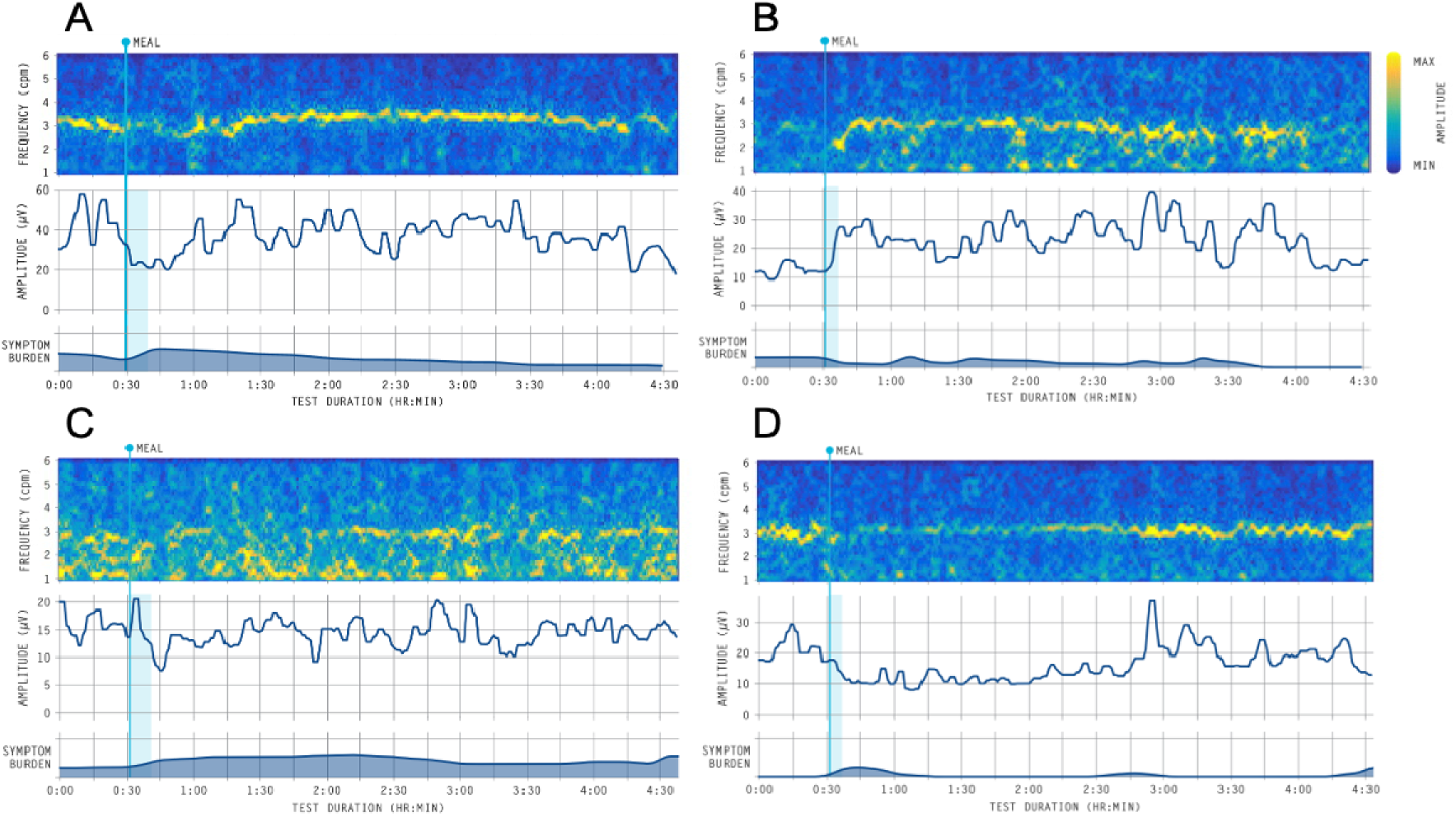
Example spectrograms, for various presentations of GERD and non-GERD. A) non-GERD patient with normal spectra. B) GERD patient with normal spectra. C) GERD patient with dysrhythmic spectral presentation. D) GERD patient demonstrating a delayed meal response.

**Table 3.**
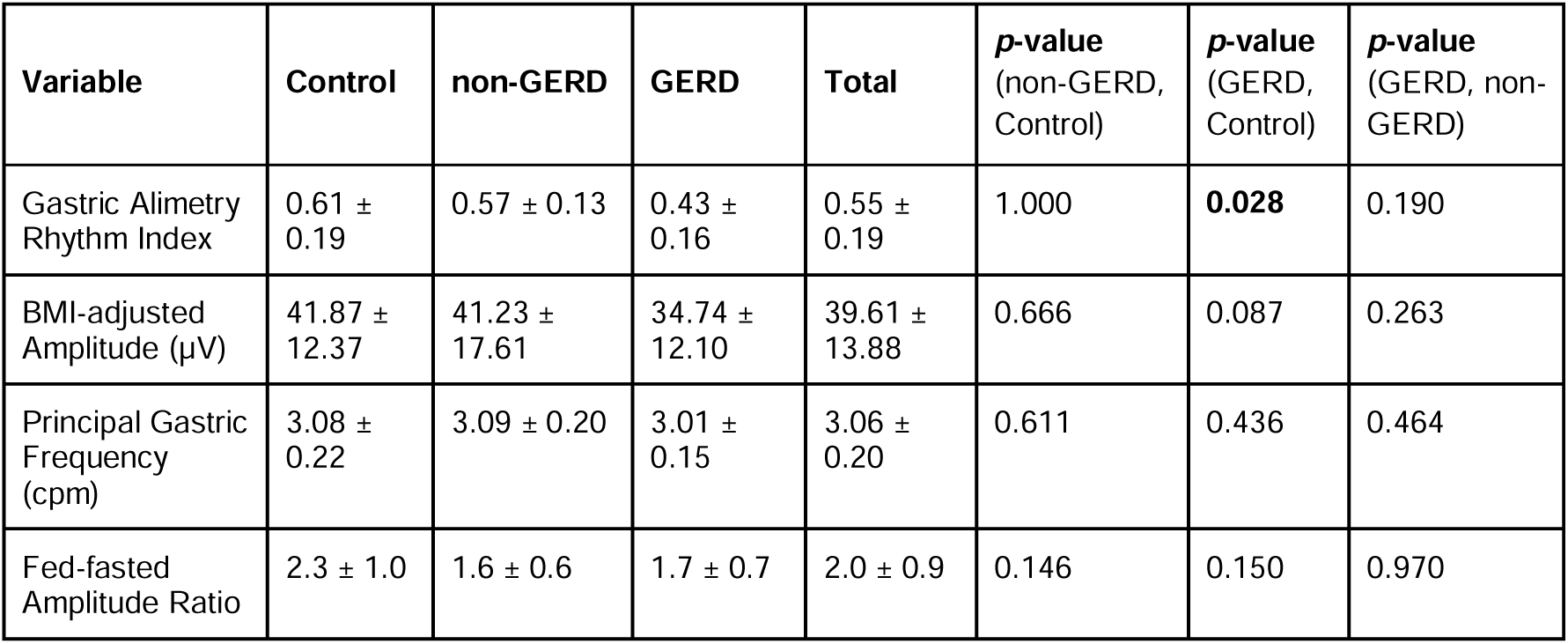
Spectral characteristics for controls and patients with and without a GERD diagnosis.

| Variable | Control | non-GERD | GERD | Total | *p*-value<br>(non-GERD,<br>Control) | *p*-value<br>(GERD,<br>Control) | *p*-value<br>(GERD, non-GERD) |
| --- | --- | --- | --- | --- | --- | --- | --- |
| Gastric Alimetry<br>Rhythm Index | 0.61 ±<br>0.19 | 0.57 ± 0.13 | 0.43 ±<br>0.16 | 0.55 ±<br>0.19 | 1.000 | **0.028** | 0.190 |
| BMI-adjusted<br>Amplitude (μV) | 41.87 ±<br>12.37 | 41.23 ±<br>17.61 | 34.74 ±<br>12.10 | 39.61 ±<br>13.88 | 0.666 | 0.087 | 0.263 |
| Principal Gastric<br>Frequency<br>(cpm) | 3.08 ±<br>0.22 | 3.09 ± 0.20 | 3.01 ±<br>0.15 | 3.06 ±<br>0.20 | 0.611 | 0.436 | 0.464 |
| Fed-fasted<br>Amplitude Ratio | 2.3 ± 1.0 | 1.6 ± 0.6 | 1.7 ± 0.7 | 2.0 ± 0.9 | 0.146 | 0.150 | 0.970 |

**Fig. 2.**
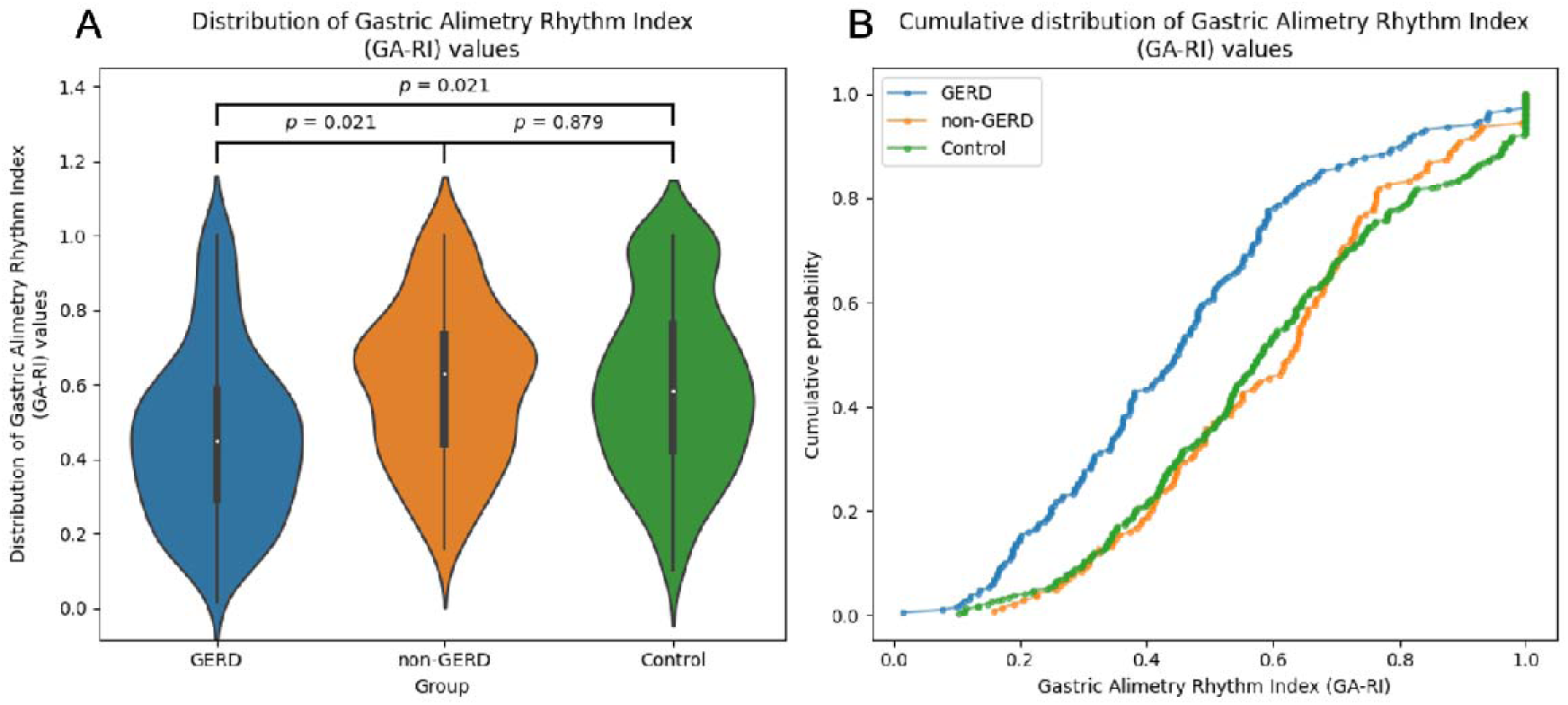
Distribution of Gastric Alimetry Rhythm Index (GA-RI) values for GERD patients, non-GERD patients, and controls. A) GERD patients had lower GA-RI values than non-GERD patients and controls. B) Cumulative distribution showing GERD patients are more likely to have lower GA-RI scores than non-GERD patients and controls.

There were no significant differences between overall PGF, BMI-adjusted amplitude, and ff-AR between groups (**Table 3**).

Both GERD and non-GERD patients displayed a ‘long-lag’ or delayed amplitude response postprandially (**Fig. 1**). This feature was quantified as decreased meal response ratio (MMR) for GERD (0.94 ± 0.19 vs. 1.37 ± 0.58; *p* = 0.006) and non-GERD (0.84 ± 0.28 vs.1.37 ± 0.58; *p* = 0.003) patients compared to controls. MRR was not statistically different between GERD and non-GERD patients (*p* = 0.379).

GA-RI was negatively correlated with symptom burden including GCSI and PAGI-SYM score, as summarised in **Fig. 3A** (all *p* < 0.05). In addition, a moderate negative correlation was identified between GA-RI and DeMeester score (N = 20, *r* = -0.45, *p* = 0.042, **Fig. 3B**), though statistical significance was lost when analysing diagnostic subgroups (GERD: n = 12, *r* = -0.36, *p* = 0.246; non-GERD: n = 8, *r* = -0.09, *p* = 0.816; **Fig. 3B**).

**Fig. 3.**
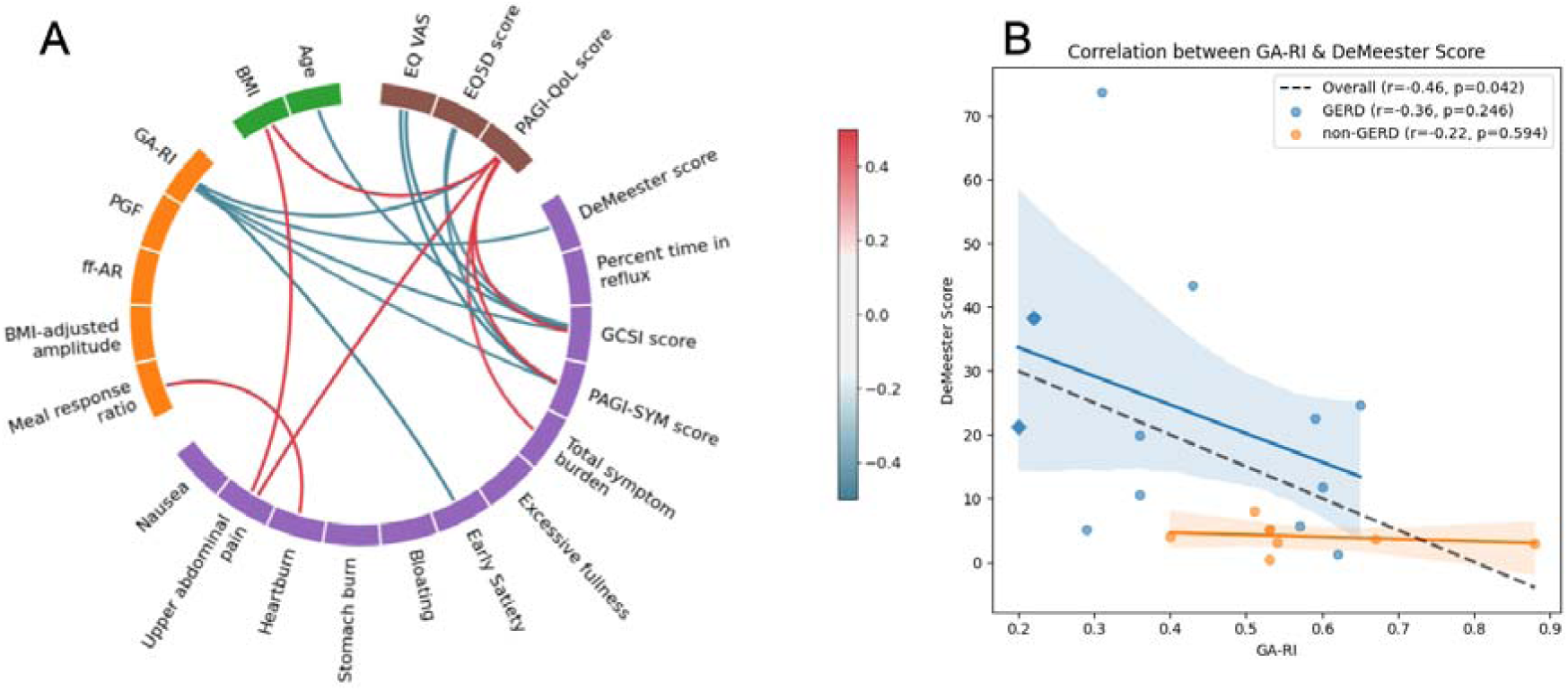
A) Wheel plot showing the relationship between BSGM metrics, patient factors, Gastric Alimetry symptom scores, and quality of life scores. Values shown are Pearson’s correlation coefficients that surpassed the significance threshold after adjustment for multiple comparisons using a Benjamini–Hochberg correction. GA-RI; Gastric Alimetry rhythm index, PGF; principal gastric frequency, ff-AR; fed:fasted amplitude ratio. B) Relationship between GA-RI (calculated over the 4 hr BSGM recording) and Overall DeMeester score (calculated across a 24 hr pH period) for GERD and non-GERD patients. GERD patients had greater average DeMeester scores (23.2 ± 20.5) than non-GERD patients (3.75 ± 2.3; *p* = 0.007). For the overall cohort there was a significant negative correlation between GA-RI and DeMeester score (*r* = -0.46, *p* = 0.042), though statistical significance was not observed for GERD patients (*r* = -0.36, *p* = 0.246) and non-GERD patients (*r* = -0.22, *p* = 0.594), when analysed separately. Diamond data points represent individuals with dysrhythmic BSGM activity.

### Time-Dependent Analysis

BSGM data and pH monitoring data were time synchronised in 15-minute epochs, and adjusted for inter-individual variations via mean-centering. In GERD patients, mean-centred changes in GA-RI were weakly correlated with time spent in reflux during the BSGM test (number of epochs = 71, *r* = 0.25, *p* = 0.038; **Fig. 4**). In contrast, this relationship was not significant in non-GERD patients (number of epochs = 14, *r* = 0.45, *p* = 0.111), nor the combined patient cohort (*n* = 85, *r* = 0.17, *p* = 0.116).

**Fig. 4.**
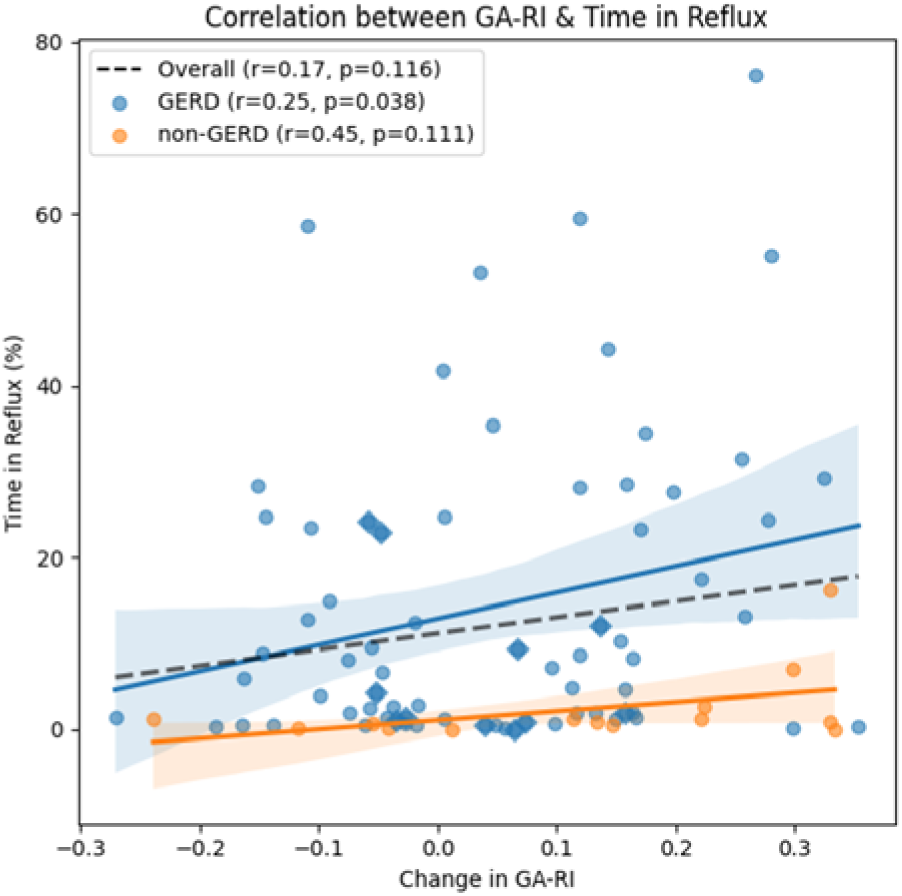
Relationship between Gastric Alimetry Rhythm Index (GA-RI) and time spent in reflux per epoch. A) Time spent in reflux expressed as the percentage of epoch duration per patient. In GERD patients, increases in rhythm stability (GA-RI) were moderately correlated increased time in reflux (*r* = 0.25, *p* = 0.038). In contrast, this relationship was not significant in non-GERD patients (*r* = 0.45, *p* = 0.111), nor was it reflected in the combined patient cohort (*r* = 0.17, *p* = 0.116). Diamond data points represent individuals with dysrhythmic BSGM activity.

Comparing mean-centred changes in GA-RI against simultaneous mean-centred changes in symptom burden revealed several relationships (**Fig. 5**). Changes in GA-RI were significantly negatively correlated with excessive fullness (*r* = -0.24, *p* < 0.001), with a stronger effect in non-GERD (*r* = -0.42, *p* < 0.001) than GERD patients (*r* = -0.17, *p* = 0.022, **Fig. 5A**). No significant association was found between GA-RI and heartburn in either patient groups (all *p* > 0.05; **Fig. 5B**). Changes in GA-RI were also weakly negatively correlated with nausea in the overall cohort (*r* = -0.21, *p* < 0.001), and in non-GERD patients (*r* = -0.37, *p* < 0.001), but not in GERD patients (*r* = -0.07, *p* = 0.341; **Fig. 5C**).

**Fig. 5.**
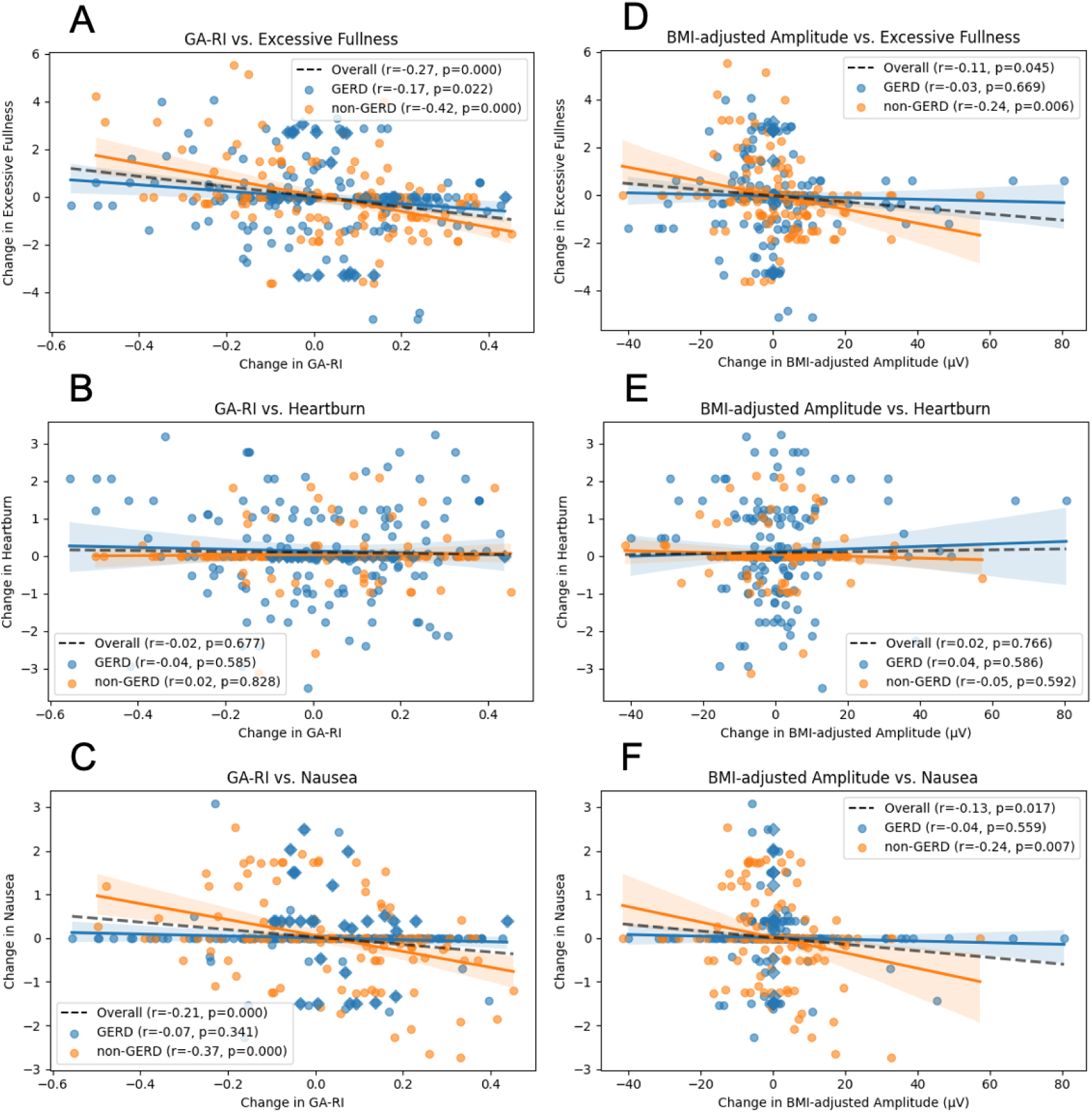
Symptomatic changes in relation to changing Gastric Alimetry Rhythm Index (GA-RI) and BMI-adjusted amplitude across all patients. A) Changes in GA-RI and excessive fullness were weakly negatively correlated in GERD patients and strongly negatively correlated in non-GERD patients. B) There was no relationship between changes in GA-RI and heartburn for either patient group. C) Changes in GA-RI and nausea were moderately negatively correlated in non-GERD patients, but there was no significance in GERD patients. D) Changes in BMI-adjusted amplitude and excessive fullness were moderately negatively correlated in non-GERD patients, but there was no significance in GERD patients. E) There was no relationship between changes in BMI-adjusted amplitude and heartburn for either patient group. F) Changes in BMI-adjusted amplitude and excessive fullness were moderately negatively correlated in non-GERD patients, but there was no significance in GERD patients. Diamond data points represent individuals with dysrhythmic BSGM activity.

To account for the potential effect of concurrent PPI use on heartburn symptoms, a sensitivity analysis excluding patients who underwent pH testing while on PPI therapy is presented in **Supplementary Fig. 3**. Results were consistent with the original analysis, except for the relationship between BMI-adjusted amplitude and excessive fullness.

Compared to the whole cohort analysis, the sensitivity analysis revealed stronger overall associations, and an emergent correlation for the GERD group between amplitude and fullness (original: *r* = -0.03, *p* = 0.669 vs. sensitivity: *r* = -0.20, *p* = 0.020).

## Discussion

This study defined the electrophysiological profile and symptom associations in patients with suspected GERD through simultaneous BSGM and pH monitoring. Reduced GA-RI (gastric rhythm stability) was associated with higher symptom burden and greater DeMeester score, suggesting that reduced gastric myoelectrical stability may increase a patient’s propensity toward reflux events. BSGM could provide novel biomarkers of symptom severity in patients with suspected GERD.

While there was an overall association between unstable gastric rhythm and greater symptom or reflux burden, there was no time-dependent correlation between rhythm instability and heartburn symptoms. Instead, transient decreases in rhythm stability were temporally associated with symptoms of excessive fullness and nausea, suggesting gastric neuromuscular dysfunction is comorbid in subsets of patients with GERD symptoms. This finding remained consistent on sensitivity analysis after excluding patients who completed tests on PPI medication.

These findings are consistent with previous studies but also significantly extend them by offering novel insights into GERD pathophysiology. Gastric dysrhythmia is temporally associated with symptoms of excessive fullness,^10^ nausea,^24^ and broadly associated with nausea and vomiting syndromes,^25^ diabetic gastropathy,^21^ and post-surgical cohorts.^26,27^ Gastric dysrhythmia is also more prevalent in patients with GERD according to legacy EGG techniques,^7^ but no temporal association with reflux has been demonstrated previously. It follows that gastric dysrhythmia may predispose patients to gastric symptoms, but may not directly generate heartburn symptoms and reflux events. In addition, the temporal correlation between gastric rhythm biomarkers, and functional dyspepsia or gastroparesis-like symptoms, highlights the close relationship between these overlapping disorders, and supports the emerging role of BSGM in determining motility contributions to symptoms.^28^

Our cohort identified abnormal overall gastric myoelectrical activity (*i.e.* outside of normative reference intervals) in 10% of all patients presenting for 24 hr pH monitoring, and in 16.7% of patients with confirmed GERD. Two GERD patients were classified as having a dysrhythmic phenotype (GA-RI < 0.25) with high DeMeester scores. Our data suggest that only a small portion of patients with GERD display major neuromuscular abnormalities and disorganised gastric activity due to depletion of stomach Interstitial Cell of Cajal networks.^29–31^ Previous data have suggested gastric dysrhythmias are more prevalent in patients who report regurgitation symptoms in addition to typical heartburn symptoms,^8^ suggesting distinct pathophysiological subgroups of patients within those reporting reflux symptoms alone.

BSGM with Gastric Alimetry® may be a diagnostic aid in patients with atypical or refractory GERD symptoms and a possible overlapping gastric rhythm disorder. Approximately 25% of patients may have unsatisfactory relief of reflux symptoms despite PPI therapy.^32,33^ Further investigations using endoscopy, manometry, pH monitoring, and gastric emptying are indicated in select cases.^33^ Our data indicates BSGM as a non-invasive method to evaluate neuromuscular dysfunction as a correlated contributor to GERD and overlapping functional dyspepsia.

Post-prandial peak gastric amplitude was delayed in both GERD and non-GERD patients undergoing 24 hr pH monitoring compared with healthy controls. This contrasts the recordings of matched controls, whose amplitude increased soon after the meal intervention. The mechanism of this behaviour is poorly understood, and further studies are needed to understand its relation to GERD.

While the presence of BSGM abnormalities in this cohort is significantly lower than the 46% prevalence of EGG abnormalities in GERD patients reported by a recent meta-analysis,^7^ this is expected because legacy EGG findings require cautious interpretation. Abnormal frequencies measured by legacy systems may include biologically implausible gastric frequencies (*e.g.*, < 1.5 or > 5 cpm) and conflate artifacts with abnormal gastric activity, limiting their modern applicability.^34^ Neuromuscular abnormalities indicated by sustained dysrhythmic gastric slow wave activity would also not necessarily be captured by traditional EGG recordings, as these patterns may occur at normogastric frequencies or are localised to distinct regions of the organ, highlighting one of the benefits of high-resolution BSGM techniques compared to legacy EGG.^15,35^

A number of patients did not withhold PPIs prior to the 24 hr pH catheter monitoring, due to a combination of clinician preference, and patient intolerability of symptoms without PPI therapy. However, sensitivity analysis excluding these patients yielded consistent results.

Furthermore, different pH measurement devices were used between centres, and unmeasured differences in device sensitivity to pH may exist. The presence of a monitoring catheter in the lower esophagus could theoretically disrupt the physiology of the stomach, potentially impacting the meal response; however, this is felt to be unlikely. Lastly, as hypothesis generating work, the patient cohort was not specifically selected. Future studies on specific subgroups of patients, such as patients with medically refractory GERD, may more clearly delineate the prevalence of myoelectrical abnormalities. Despite these limitations, this study provides significant insight into the pathophysiology of GERD by using a novel non-invasive diagnostic technique and symptom logging simultaneously with validated pH monitoring. Further studies to clarify the impact of presence of distal esophageal catheters on delayed amplitude responses, the role of retrograde slow wave activity on GERD, and on specific patients with medication-responsive, and medication-refractory cases are needed.

In conclusion, gastric rhythm instability is associated with both increased symptom severity and overall acid exposure in GERD patients. As such, GA-RI is an emerging biomarker for disease severity in these patients. While no temporal link between periods of reduced GA-RI and heartburn was found, rhythm instability was temporally associated with increased nausea and fullness. These findings suggest that gastric dysrhythmia may increase the propensity for gastroesophageal reflux, but is not a causative mechanism.

## Conflicts of interest

Greg O’Grady and Armen Gharibans hold grants and intellectual property in the field of gastrointestinal electrophysiology and are the co-founders at Alimetry Ltd.

William Xu, Sam Simmonds, Daphne Foong, Chris Varghese, Christopher N Andrews, Gabe Schamberg, Armen Gharibans, Stefan Calder are members of Alimetry Ltd.

The remaining authors have no conflicts of interest to declare.

## Author contributions

Planning and conduction of study: WX, SB, CV, CNA, AG, TLA, DR, VH, SC, GOG

Collecting data: WX, DF, DR, VH, SC, GOG

Data analysis: WX, SS, GB, AG, GOG

Manuscript writing and review: All authors

All authors have reviewed and approved the final draft submitted.

## Data Availability

All data produced in the present study are available upon reasonable request to the authors

## Supplementary materials

### Supplementary tables and figures

**Supplementary Table 1.**
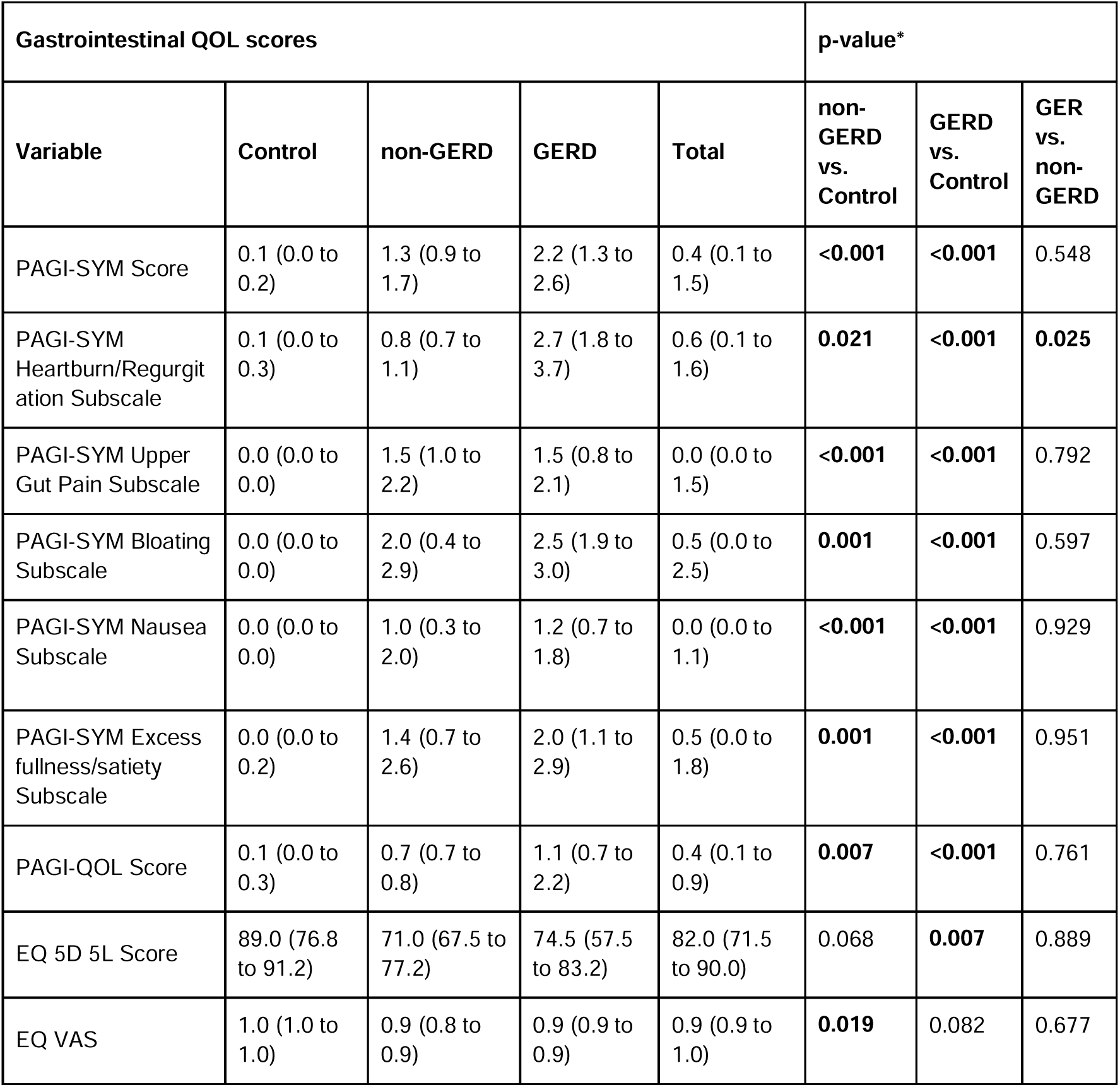
Baseline symptom, quality of life, and BSGM metric data between controls and patients undergoing pH testing with or without evidence of gastro-oesophageal reflux disease. PAGI-SYM scores are based on self-reported symptoms of the previous 2 weeks. Bolded values indicate *p*-values below the significance threshold (*p* < 0.05), following a post-hoc Tukey test with Benjamini-Hochberg’s corrections for multiple comparisons.

**Supplementary Table 2:**
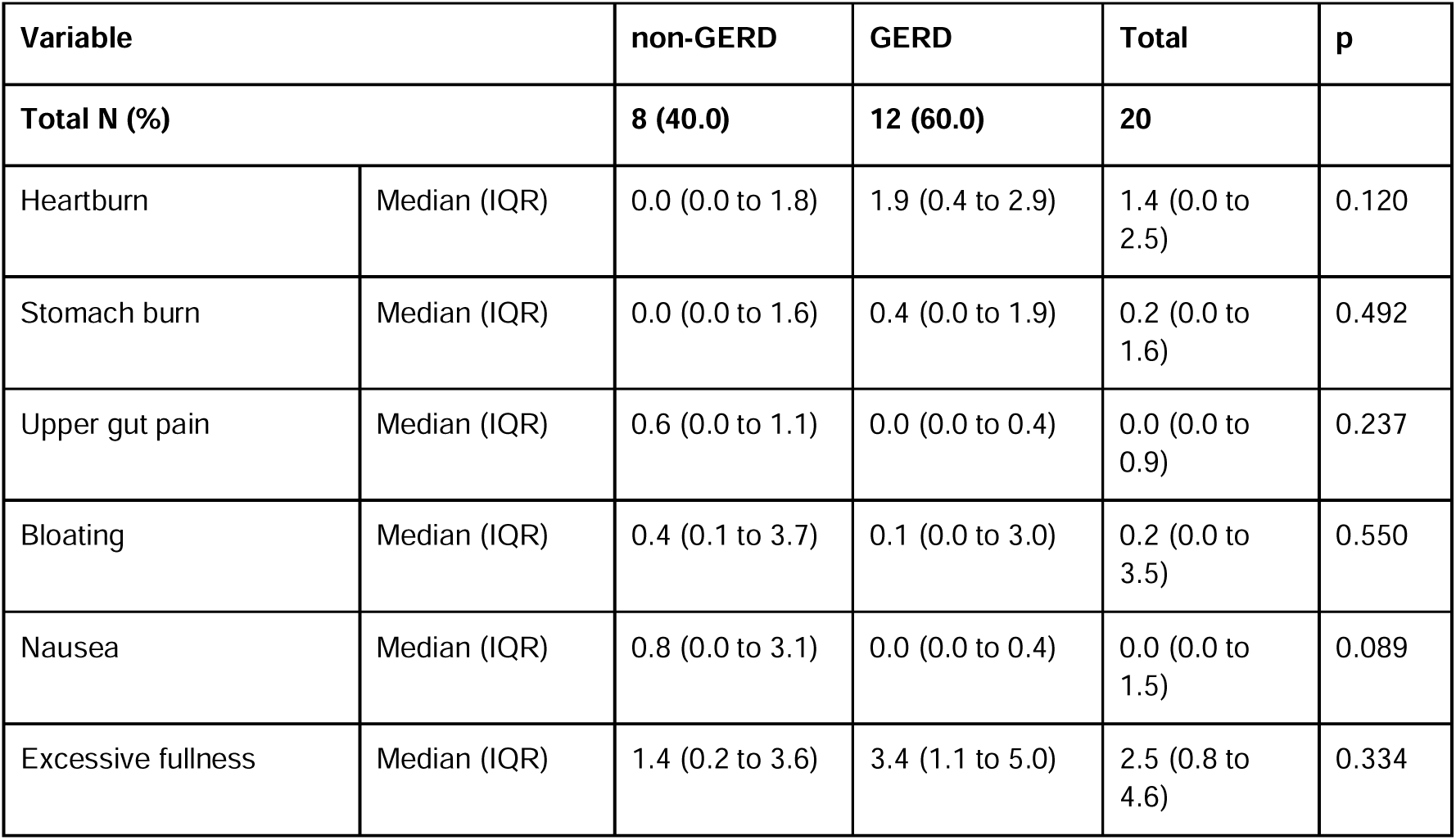
Symptom scores are based on symptom logging completed during simultaneous pH impedance and Gastric Alimetry testing.

**Supplementary Fig. 1:**
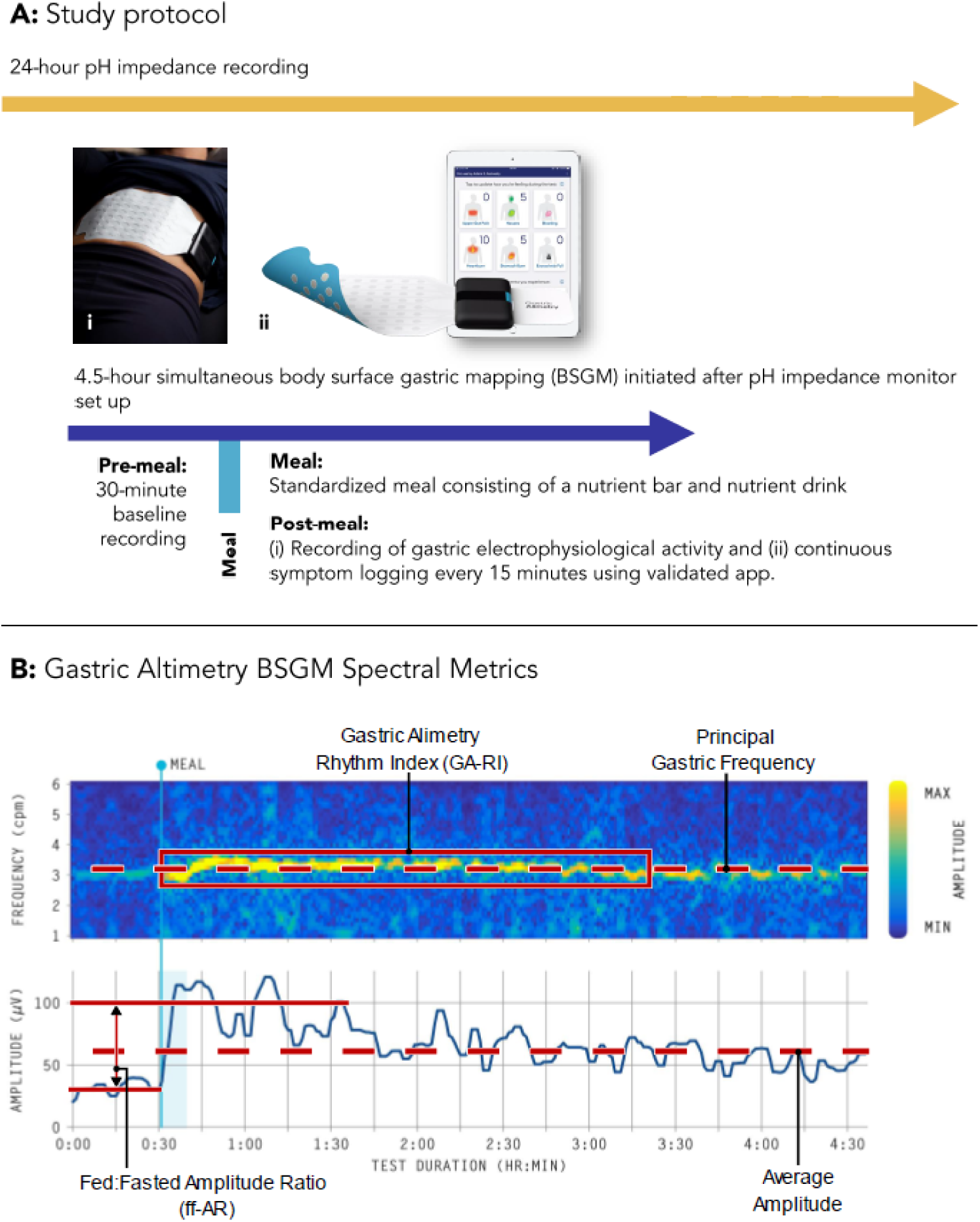
A) Study protocol for simultaneous pH monitoring and Gastric Alimetry body surface gastric mapping (BSGM). B) Example BSGM spectra (top), amplitude plot (bottom), and quantified metrics (Gastric Alimetry Rhythm Index, principal gastric frequency, fed:fasted amplitude ratio, and average BMI-adjusted amplitude) for a healthy control.

**Supplementary Fig. 2.**
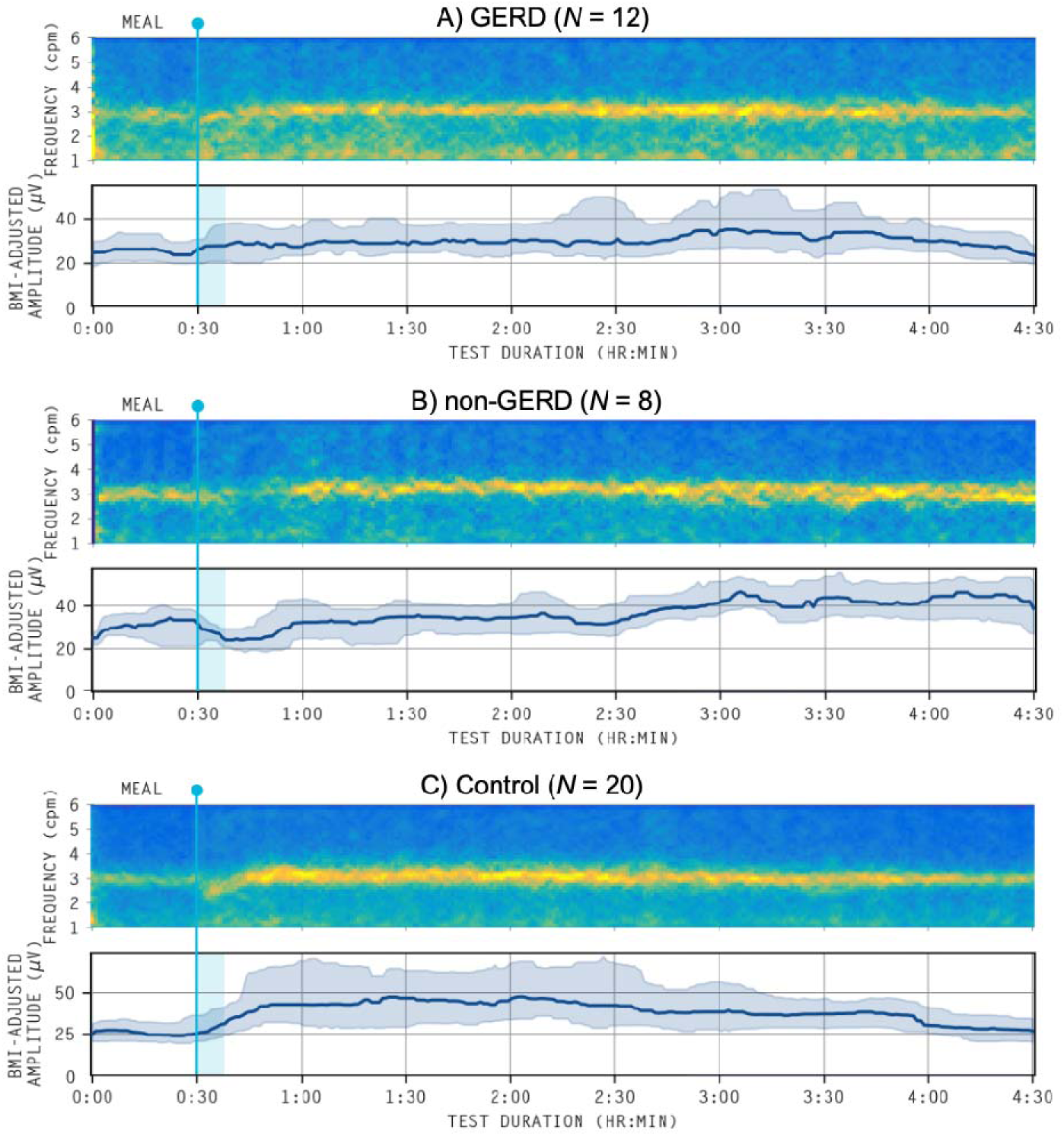
Normalised average spectrograms for each group. A) GERD patients, B) non-GERD patients, and C) controls. Post-prandially, controls had an increase in BMI-adjusted amplitude, while GERD and non-GERD patients had lagged amplitude responses.

**Supplementary Fig. 3.**
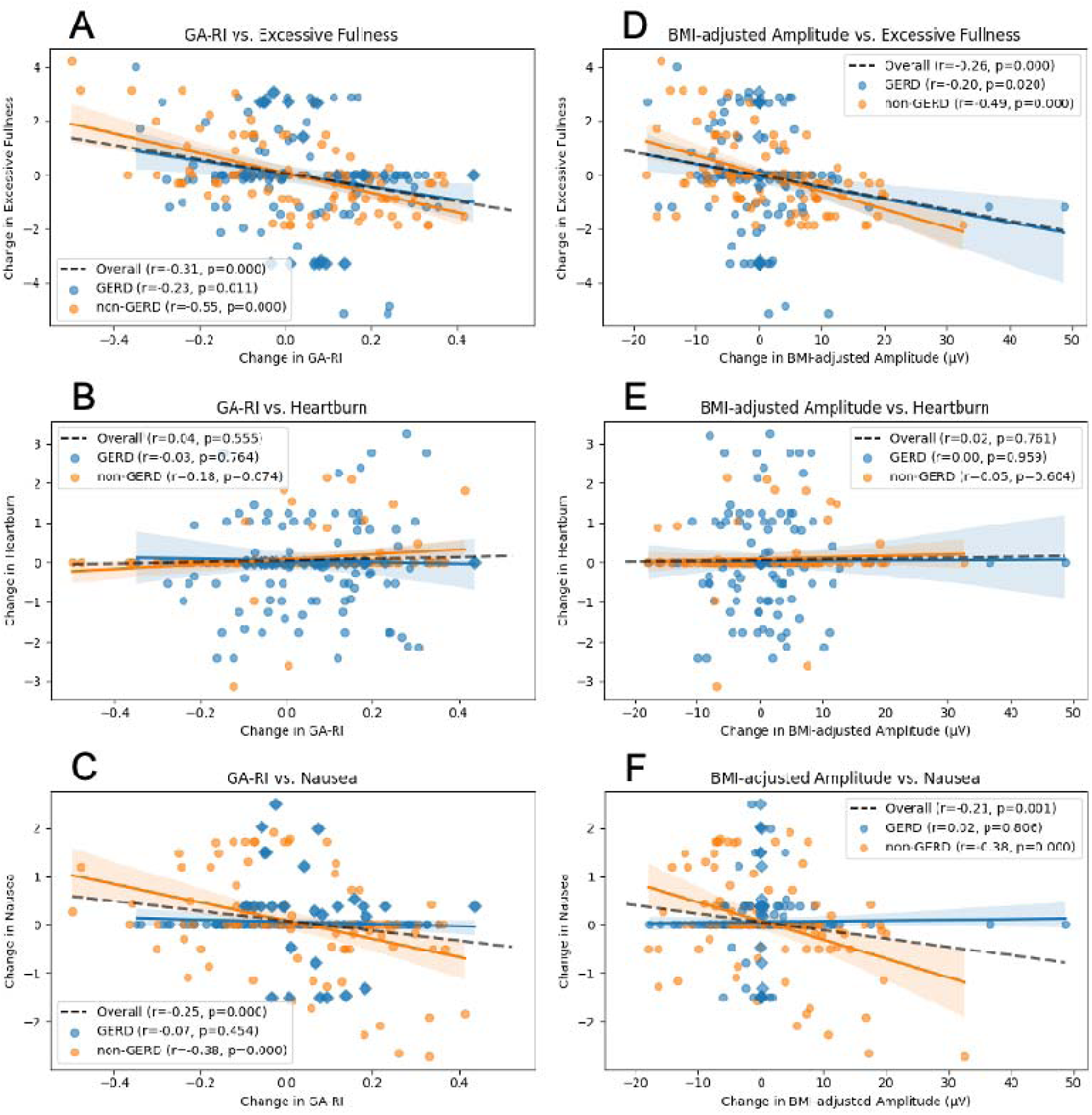
Sensitivity analysis for **Fig. 5**. Symptomatic changes in relation to changing Gastric Alimetry Rhythm Index (GA-RI) and BMI-adjusted amplitude across patients who had their PPI withheld prior to the study (*n* = 15). This sensitivity analysis revealed a stronger association between BMI-adjusted amplitude and excessive fullness and an emergent correlation for the GERD group (*r* = - 0.03, *p* = 0.669 vs. *r* = -0.20, *p* = 0.020). There were no other differences between these 15 patients and the overall cohort (*N* = 20). A) Changes in GA-RI and excessive fullness were weakly negatively correlated in GERD patients and strongly negatively correlated in non-GERD patients. B) There was no relationship between changes in GA-RI and heartburn for either patient group. C) Changes in GA-RI and nausea were moderately negatively correlated in non-GERD patients, but there was no significance in GERD patients. D) Changes in BMI-adjusted amplitude and excessive fullness were moderately negatively correlated in both patient groups. E) There was no relationship between changes in BMI-adjusted amplitude and heartburn for either patient group. F) Changes in BMI-adjusted amplitude and excessive fullness were moderately negatively correlated in non-GERD patients, but there was no significance in GERD patients. Diamond data points represent individuals with dysrhythmic BSGM activity.

