## Supplementary Figures for "Association between gastric rhythm and gastroesophageal reflux defined by simultaneous body surface gastric mapping and 24-hour pH testing"

### Supplementary materials

**Supplementary Table 1*.*** Baseline symptom, quality of life, and BSGM metric data between controls and patients undergoing pH testing with or without evidence of gastro-oesophageal reflux disease. PAGI-SYM scores are based on self-reported symptoms of the previous 2 weeks. Bolded values indicate *p*-values below the significance threshold (*p* < 0.05), following a post-hoc Tukey test with Benjamini-Hochberg’s corrections for multiple comparisons.

| **Gastrointestinal QOL scores** | | | | | **p-value*** | | |
| --- | --- | --- | --- | --- | --- | --- | --- |
| **Variable** | **Control** | **non-GERD** | **GERD** | **Total** | **non- GERD vs. Control** | **GERD vs. Control** | **GER vs. non- GERD** |
| PAGI-SYM Score | 0.1 (0.0 to 0.2) | 1.3 (0.9 to 1.7) | 2.2 (1.3 to 2.6) | 0.4 (0.1 to 1.5) | **<0.001** | **<0.001** | 0.548 |
| PAGI-SYM Heartburn/Regurgitation Subscale | 0.1 (0.0 to 0.3) | 0.8 (0.7 to 1.1) | 2.7 (1.8 to 3.7) | 0.6 (0.1 to 1.6) | **0.021** | **<0.001** | **0.025** |
| PAGI-SYM Upper Gut Pain Subscale | 0.0 (0.0 to 0.0) | 1.5 (1.0 to 2.2) | 1.5 (0.8 to 2.1) | 0.0 (0.0 to 1.5) | **<0.001** | **<0.001** | 0.792 |
| PAGI-SYM Bloating Subscale | 0.0 (0.0 to 0.0) | 2.0 (0.4 to 2.9) | 2.5 (1.9 to 3.0) | 0.5 (0.0 to 2.5) | **0.001** | **<0.001** | 0.597 |
| PAGI-SYM Nausea Subscale | 0.0 (0.0 to 0.0) | 1.0 (0.3 to 2.0) | 1.2 (0.7 to 1.8) | 0.0 (0.0 to 1.1) | **<0.001** | **<0.001** | 0.929 |
| PAGI-SYM Excess fullness/satiety Subscale | 0.0 (0.0 to 0.2) | 1.4 (0.7 to 2.6) | 2.0 (1.1 to 2.9) | 0.5 (0.0 to 1.8) | **0.001** | **<0.001** | 0.951 |
| PAGI-QOL Score | 0.1 (0.0 to 0.3) | 0.7 (0.7 to 0.8) | 1.1 (0.7 to 2.2) | 0.4 (0.1 to 0.9) | **0.007** | **<0.001** | 0.761 |
| EQ 5D 5L Score | 89.0 (76.8 to 91.2) | 71.0 (67.5 to 77.2) | 74.5 (57.5 to 83.2) | 82.0 (71.5 to 90.0) | 0.068 | **0.007** | 0.889 |
| EQ VAS | 1.0 (1.0 to 1.0) | 0.9 (0.8 to 0.9) | 0.9 (0.9 to 0.9) | 0.9 (0.9 to 1.0) | **0.019** | 0.082 | 0.677 |

**Supplementary Table 2:** Symptom scores are based on symptom logging completed during simultaneous pH impedance and Gastric Alimetry testing.

| **Variable** | | **non-GERD** | **GERD** | **Total** | **p** |
| --- | --- | --- | --- | --- | --- |
| **Total N (%)** | | **8 (40.0)** | **12 (60.0)** | **20** |  |
| Heartburn | Median (IQR) | 0.0 (0.0 to 1.8) | 1.9 (0.4 to 2.9) | 1.4 (0.0 to 2.5) | 0.120 |
| Stomach burn | Median (IQR) | 0.0 (0.0 to 1.6) | 0.4 (0.0 to 1.9) | 0.2 (0.0 to 1.6) | 0.492 |
| Upper gut pain | Median (IQR) | 0.6 (0.0 to 1.1) | 0.0 (0.0 to 0.4) | 0.0 (0.0 to 0.9) | 0.237 |
| Bloating | Median (IQR) | 0.4 (0.1 to 3.7) | 0.1 (0.0 to 3.0) | 0.2 (0.0 to 3.5) | 0.550 |
| Nausea | Median (IQR) | 0.8 (0.0 to 3.1) | 0.0 (0.0 to 0.4) | 0.0 (0.0 to 1.5) | 0.089 |
| Excessive fullness | Median (IQR) | 1.4 (0.2 to 3.6) | 3.4 (1.1 to 5.0) | 2.5 (0.8 to 4.6) | 0.334 |

**Supplementary Fig. 1**: A) Study protocol for simultaneous pH monitoring and Gastric Alimetry body surface gastric mapping (BSGM). B) Example BSGM spectra (top), amplitude plot (bottom), and quantified metrics (Gastric Alimetry Rhythm Index, principal gastric frequency, fed:fasted amplitude ratio, and average BMI-adjusted amplitude) for a healthy control.

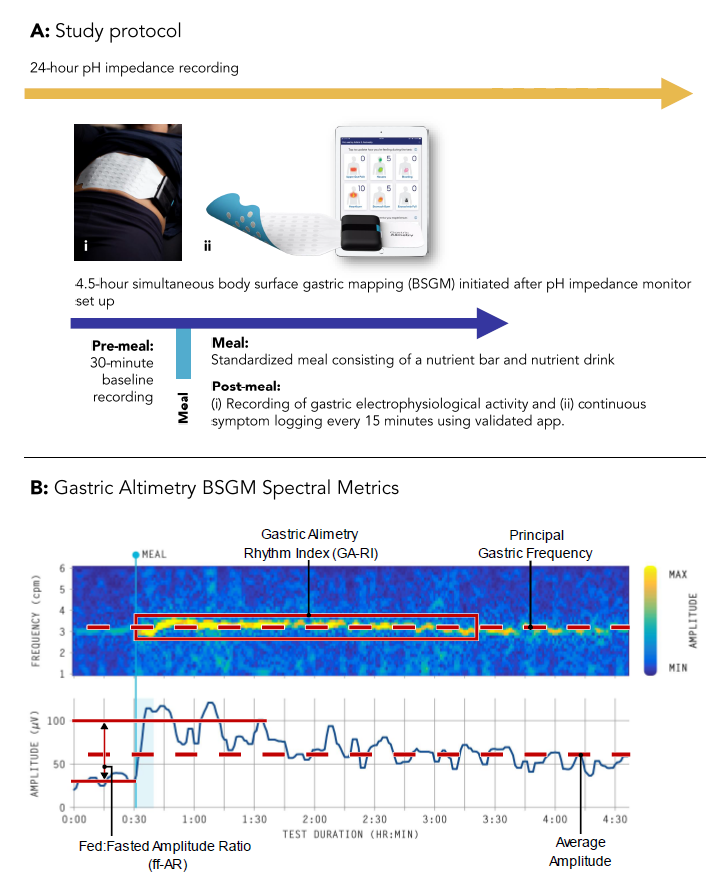

**Supplementary Fig. 2**. Normalised average spectrograms for each group. A) GERD patients, B) non-GERD patients, and C) controls. Post-prandially, controls had an increase in BMI-adjusted amplitude, while GERD and non-GERD patients had lagged amplitude responses.

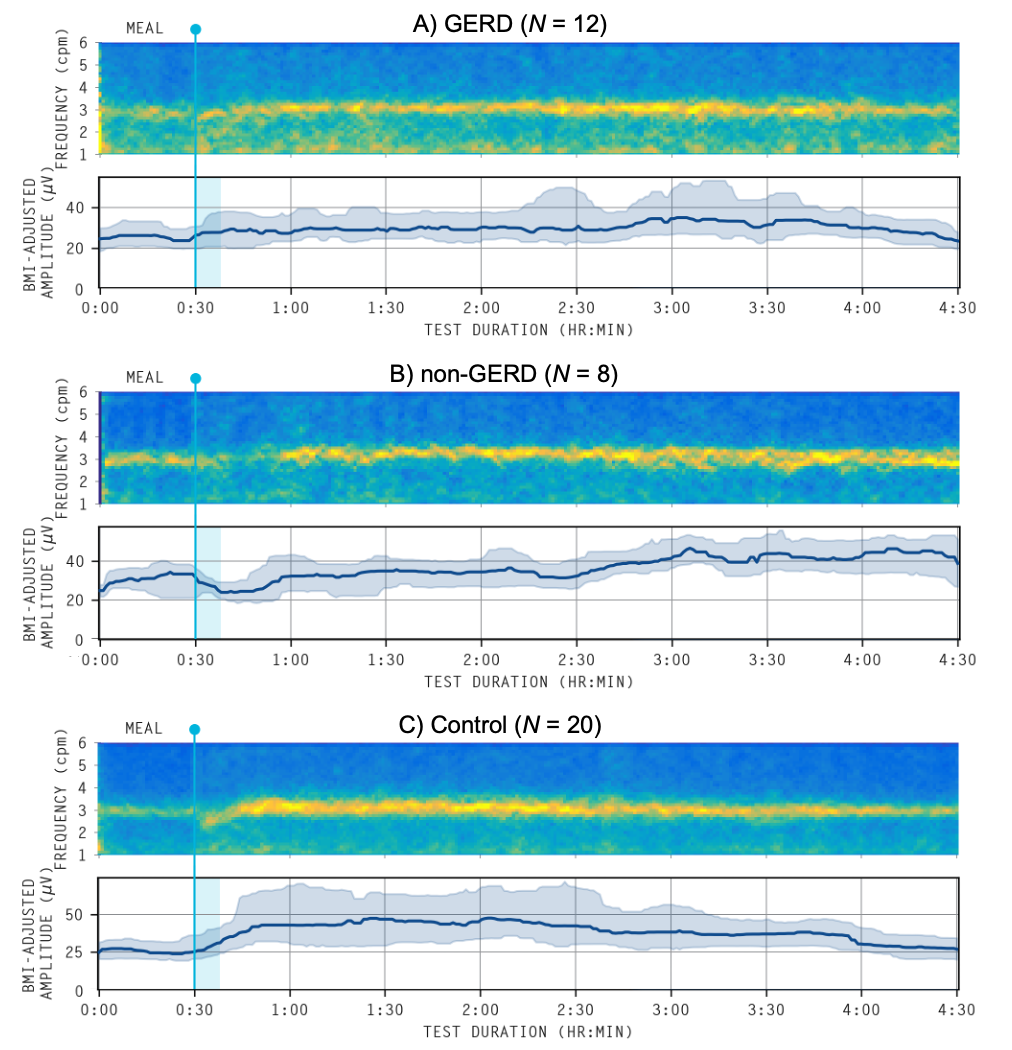

**Supplementary Fig. 3.** Sensitivity analysis for Fig. 5. Symptomatic changes in relation to changing Gastric Alimetry Rhythm Index (GA-RI) and BMI-adjusted amplitude across patients who had their PPI withheld prior to the study (*n* = 15). This sensitivity analysis revealed a stronger association between BMI-adjusted amplitude and excessive fullness and an emergent correlation for the GERD group (*r* = -0.03, *p* = 0.669 vs. *r* = -0.20, *p* = 0.020). There were no other differences between these 15 patients and the overall cohort (*N* = 20). A) Changes in GA-RI and excessive fullness were weakly negatively correlated in GERD patients and strongly negatively correlated in non-GERD patients. B) There was no relationship between changes in GA-RI and heartburn for either patient group. C) Changes in GA-RI and nausea were moderately negatively correlated in non-GERD patients, but there was no significance in GERD patients. D) Changes in BMI-adjusted amplitude and excessive fullness were moderately negatively correlated in both patient groups. E) There was no relationship between changes in BMI-adjusted amplitude and heartburn for either patient group. F) Changes in BMI-adjusted amplitude and excessive fullness were moderately negatively correlated in non-GERD patients, but there was no significance in GERD patients. Diamond data points represent individuals with dysrhythmic BSGM activity.

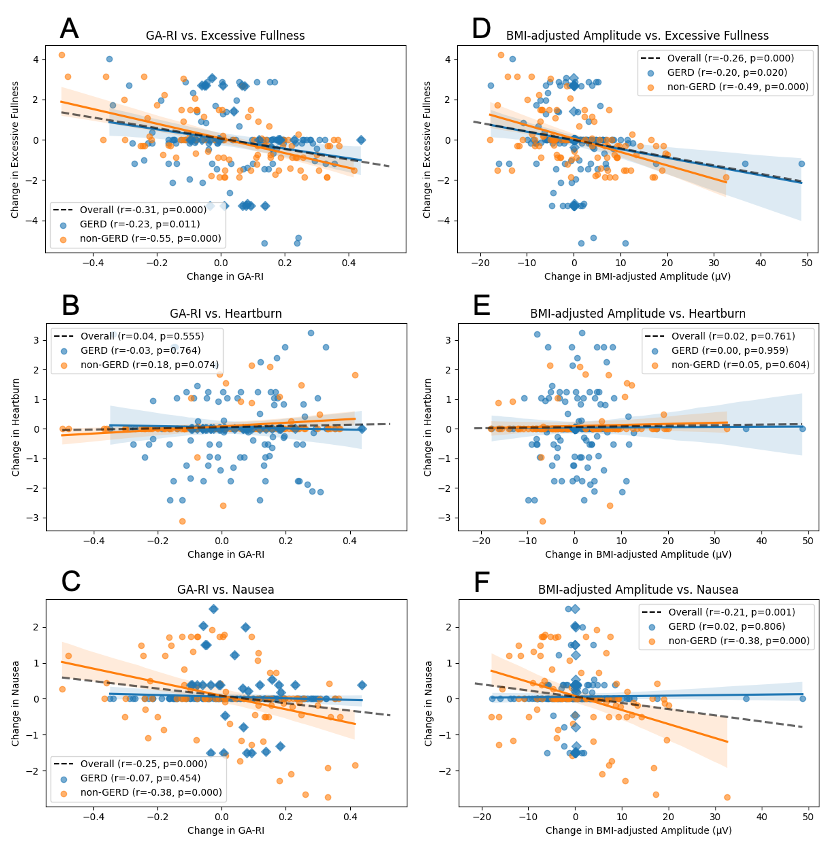
